# Why more doctors may not mean more essential-specialty physicians

**DOI:** 10.64898/2025.12.25.25343018

**Authors:** Hojun Yu

## Abstract

**Purpose:** Korea is expanding medical school admissions on the assumption that a larger physician pool will relieve shortages in essential specialties. This article argues that the crisis is primarily one of allocation rather than absolute physician numbers, and asks whether increasing total supply will increase the number of physicians actively practicing in essential specialties.

**Current concepts:** A physician’s field of practice is not fixed at licensure. Published reanalyses of claims data indicate that many board-certified specialists in primary care display a practice department other than the specialty they trained in. As a thought experiment, entry into and retention in an essential specialty can be framed as an expected-value comparison weighing a gain term (career success and net return) against a cost term (litigation and licensing risk). This framing makes explicit which term policy affects; the parameter values are stated assumptions, not empirical estimates. Under a fixed insurance budget, added supply dilutes the return term without increasing reimbursed essential work, whereas the cost term is set by the medico-legal environment rather than supply. Displaced specialists may accumulate in substitutable aesthetic and general-practice markets, while non-substitutable essential capacity remains unfilled.

**Discussion and conclusion:** Essential-specialty participation may track specialty-specific incentives more closely than physician headcount. If so, expanding admissions while expected value remains low would not necessarily increase the essential-specialty workforce, whereas restoring returns and lowering medico-legal costs could create conditions for re-entry. Quota adjustment alone may be an incomplete policy lever. This is a conceptual, hypothesis-generating argument requiring examination with longitudinal data.

## Introduction

Korea is expanding medical school admissions on the premise that a larger physician pool will, over time, flow into the fields now experiencing the most visible shortages: cardiothoracic surgery, obstetrics and gynecology, pediatrics, general surgery, emergency medicine, and acute internal medicine. This premise treats the essential-care crisis as a problem of absolute numbers. I argue that the crisis is better explained as a problem of allocation, and that under the incentives physicians actually face, increasing total supply may leave the number of physicians actually practicing in essential specialties unchanged, or may even reduce it. I call this the paradox of expansion. The argument is conceptual: it is not a forecast derived from validated data but a proposal to reframe how this policy is reasoned about. The figures presented below are illustrative, intended to make the argument concrete rather than to measure it.

### Mismatch between licensure and actual field of practice

The starting observation is that a physician’s choice of where to practice is not fixed at the point of licensure. A board-certified specialist may practice outside the field in which they trained. A reanalysis of health insurance claims data released by the office of Rep. Shin Hyun-young of the 21st National Assembly Health and Welfare Committee indicates the scale of this (Table 1) [1]. The nature and limitations of these data should be stated first. First, they are cross-sectional as of March 2023 and are not a time series constructed under a consistent definition; they therefore do not speak directly to trends. Second, “mismatch” here refers to cases in primary care settings in which the displayed practice department differs from the physician’s board-certified specialty; specialists who continue to practice within their specialty at tertiary and general hospitals are not included in the denominator. These values therefore do not represent the proportion of all specialists in each field who have left specialty practice. Third, the displayed practice department is an item reported by the institution and may not fully correspond to the actual content of care; physicians who combine specialty practice with other practice are not distinguished. Fourth, the data were released through the press from materials submitted to the National Assembly, so the extraction criteria of the original dataset cannot be independently verified. Given these constraints, the individual values in Table 1 should be read not as precise estimates but as reference values indicating the approximate scale of the allocation problem. Even so, the fact that in primary care settings the majority of cardiothoracic surgeons and about half of general surgeons display a department other than their specialty shows that the workforce counted as trained and licensed may differ from the workforce actually delivering care in that field. Pushing more trainees into the front of the pipeline does not address why a substantial number leave essential fields at the back of it. This mismatch is not a marginal curiosity; it is concentrated in precisely the specialties the expansion policy is meant to rescue, and the persistence of essential-care avoidance over recent years has been noted repeatedly [2].

**Table 1.**
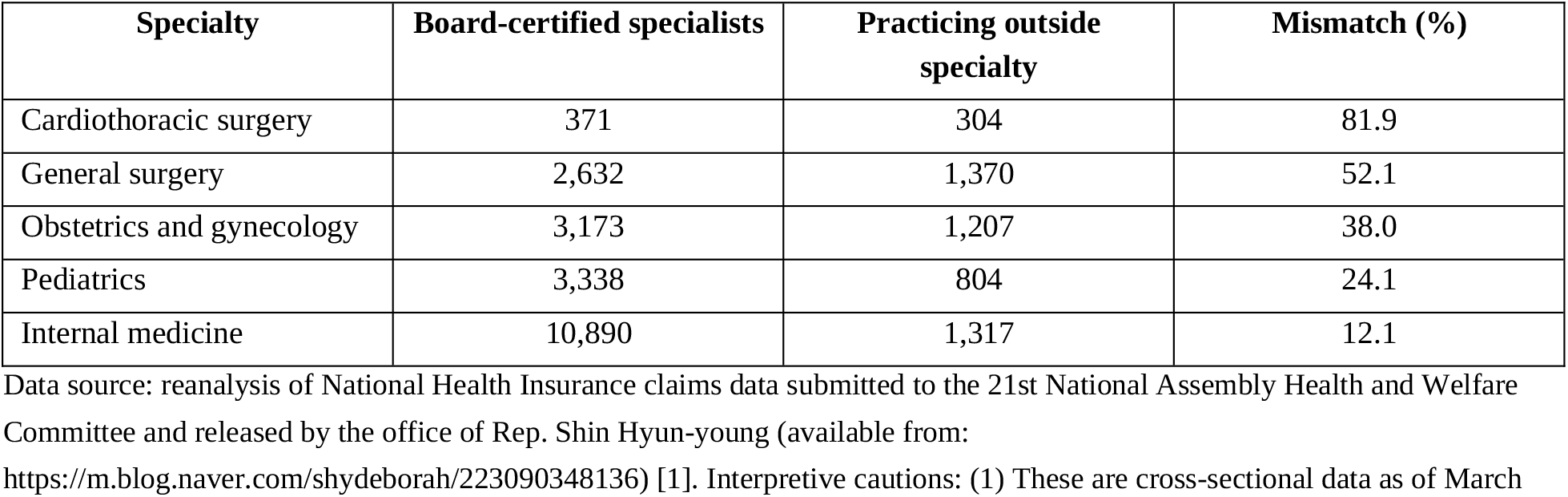
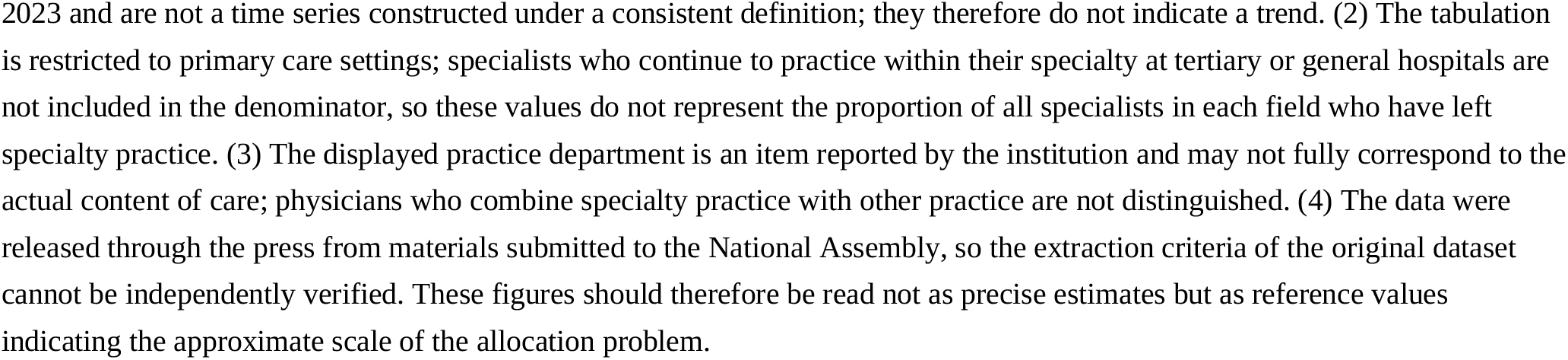
Specialists in primary care settings whose displayed practice department differs from their board-certified specialty, by essential specialty (cross-sectional data, March 2023)

| Specialty | Board-certified specialists | Practicing outside specialty | Mismatch (%) |
| --- | --- | --- | --- |
| Cardiothoracic surgery | 371 | 304 | 81.9 |
| General surgery | 2,632 | 1,370 | 52.1 |
| Obstetrics and gynecology | 3,173 | 1,207 | 38.0 |
| Pediatrics | 3,338 | 804 | 24.1 |
| Internal medicine | 10,890 | 1,317 | 12.1 |

Data source: reanalysis of National Health Insurance claims data submitted to the 21st National Assembly Health and Welfare Committee and released by the office of Rep. Shin Hyun-young (available from:
https://m.blog.naver.com/shydeborah/223090348136) [1]. Interpretive cautions: (1) These are cross-sectional data as of March
2023 and are not a time series constructed under a consistent definition; they therefore do not indicate a trend. (2) The tabulation is restricted to primary care settings; specialists who continue to practice within their specialty at tertiary or general hospitals are not included in the denominator, so these values do not represent the proportion of all specialists in each field who have left specialty practice. (3) The displayed practice department is an item reported by the institution and may not fully correspond to the actual content of care; physicians who combine specialty practice with other practice are not distinguished. (4) The data were released through the press from materials submitted to the National Assembly, so the extraction criteria of the original dataset cannot be independently verified. These figures should therefore be read not as precise estimates but as reference values indicating the approximate scale of the allocation problem.

The gradient across specialties is itself informative. The mismatch is not uniform: it is most severe in cardiothoracic surgery, followed by general surgery, obstetrics and gynecology, pediatrics, and internal medicine (Table 1). This ordering does not conflict with the risk-and-return profile of each field but rather runs parallel to it. Specialties known for high acuity, greater medico-legal exposure, and less favorable reimbursement show higher mismatch rates. This observation, however, is a correspondence found in cross-sectional data and does not demonstrate that the incentive structure caused specialty attrition. That the two patterns appear together is best understood as a reason to examine the problem in terms of the incentives each field offers rather than the absolute number of physicians.

### Specialty choice in essential medicine viewed as an expected value

Why physicians leave can be expressed as a thought experiment comparing expected values. The decision to enter an essential specialty and to remain in it is placed as a problem of weighing an expected gain against an expected cost. The gain is the probability of sustaining a successful practice career multiplied by the lifetime net return of that field; the cost is the probability of facing litigation or licensing jeopardy multiplied by what such an event costs the physician. I write this as E(V) = (P_success × R_net) − (P_suit × C_risk). This expression is not a model for measuring or predicting decisions. It is a descriptive device that makes explicit the terms a rational, risk-aware physician weighs, and that shows which of those terms current policy moves. The values entering each term are not estimated from observed data but are assumptions adopted for discussion, and no claim is made that actual physicians’ choices follow this expression. Defined this way, P_suit is best read not as the probability of losing an individual case but as the expected burden of litigation over a practice lifetime. This includes the interruption of work and the legal costs that arise even when the physician is ultimately cleared.

### A fixed budget and a cost of risk that is independent of supply

Seen this way, the number of physicians is close to irrelevant to the sign of E(V). What matters is how the essential fields stand in relation to their alternatives. When the National Health Insurance budget is effectively fixed, adding physicians dilutes the net return per physician without increasing the total volume of reimbursed essential work. The binding constraint is the revenue pool, not the number of patients. This dilution compounds a slower erosion. Under the negotiated conversion factor, the unit price of a reimbursed act has risen by roughly 2% per year, which falls short of price and wage growth, so the real value of the same work has declined for years [3]. Physicians absorb this through volume: shortening consultation time and shifting from cognitive, high-acuity care toward better-compensated imaging and laboratory work [4]. This escape route is open to routine practice but closed to the essential specialties, whose procedures and operations cannot be shortened or multiplied to match compensation. The growth of total health expenditure therefore reflects volume, population aging, and the introduction of new technologies; it does not mean that the profitability of essential work has improved.

The cost term, meanwhile, is determined by the medico-legal environment rather than by supply. Korea pursues and penalizes adverse clinical outcomes through criminal law with a severity that has few parallels among comparable health systems, and a single catastrophic judgment or license suspension can end a surgeon’s career. A rational physician weighing a high and rising cost term against a diluted and capped gain term does not leave medicine. They move to fields where the cost term is low and the return is preserved: the elastic and largely non-reimbursed aesthetic and non-covered care market. The severity of this cost term is not an exaggeration. Among comparable health systems, Korea is unusual in handling adverse outcomes through criminal courts rather than through civil or no-fault compensation channels. The probability of criminal prosecution for an adverse outcome is markedly higher than in Japan or France [5], and a criminal record or license suspension forecloses a career in a way that a civil settlement does not [6]. This asymmetry between fields in which an error becomes a lawsuit and fields in which an error ends in a refund is precisely what the cost term is meant to capture.

### Substitutability and unrealized human capital

The training capital that a mismatched specialist carries into that market is not destroyed. Neither, however, is it realized as essential-care capacity (Figure 1). What is lost is not the physician’s general clinical output—which remains useful—but the specific, non-substitutable capacity for which no alternative labor pool exists, such as cardiac surgery, obstetric delivery, and neonatal intensive care. What Korea lacks is not general outpatient care but high-risk, specialized capacity, and it is that capacity which quietly disappears through specialty mismatch.

**Figure 1.**
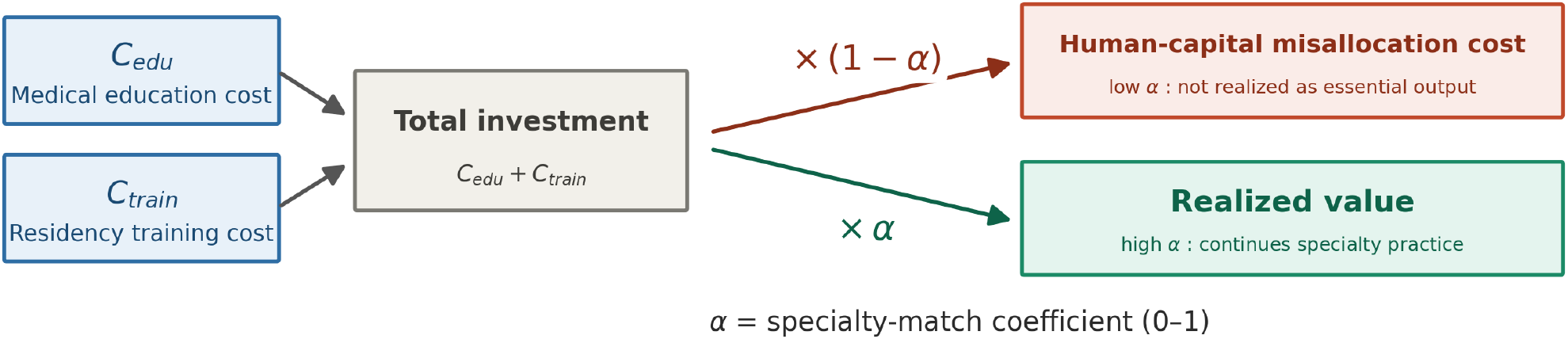
Conceptual diagram showing that human capital accumulated through training is realized as essential-specialty output only in proportion to the specialty-match coefficient α. Realized value = total investment × α, and human-capital misallocation cost = total investment × (1 − α); the two shares are complementary and sum to the total investment. When a specialist practices outside the trained field (low α), much of the investment is not realized as essential-care capacity, and this unrealized share is greater in fields with no alternative labor pool. α is a conceptual coefficient introduced for explanation and is not an observed value.

This is where comparison with a saturation argument misleads. The objection that new supply will eventually meet genuine unmet need is plausible. That logic holds for substitutable, demand-elastic services, and it is exactly there that displaced specialists accumulate. It does not hold for non-substitutable essential capacity. If cardiac surgery is reimbursed below cost, carries ruinous legal exposure, and does not even generate enough cases to maintain procedural proficiency, then no matter how many graduates are added, no surgeon will perform it. The unmet need simply remains alongside an emptied field. Counting the aesthetic market and the essential-care vacancy as a single labor pool of “physicians” compares unlike things. It sets an abundant, elastic supply of one kind of work against a shortage of an entirely different kind that the abundance can never fill.

A skeptical reader may reply that this is merely a temporary disequilibrium: as essential fields empty, scarcity should raise the reward for those who remain, drawing entrants back until the market reaches equilibrium. That is precisely the correction a classical economic model predicts, and precisely the process the current structure blocks. When reimbursement is set administratively rather than by scarcity [7], and when the dominant cost is a criminal and licensing risk that no individual physician can price into a fee, the supply that should refill the field never arrives. An emptied field is not a market moving toward equilibrium. It is a market in which the equilibrating mechanism itself has been switched off.

### The direction in which expansion moves the expected value

The direction in which expansion pushes E(V) is therefore conditional, and worth stating precisely (Table 2, Figure 2). Income dilution alone lowers the gain term, but that by itself does not necessarily make the sign of E(V) negative. With the illustrative parameters, the post-expansion value remains positive if litigation risk is held constant. The sign changes only when expansion also raises the cost term, for instance when supervised cases, experienced faculty, and procedural facilities do not grow in step with the enrollment increase and training quality declines. Given the current state of educational infrastructure in Korea, this is an assumption worth examining. Discussions of medical school admissions policy have raised the concern that a rapid increase in admissions without sufficient evidence or consultation may lead to disruption in medical education [8], and survey data from regional national university medical schools have reported shortages of lecture rooms and laboratories, faculty, and cadavers for basic practical training [9]. The educational capacity of tertiary and university hospitals may not expand sufficiently within the time frame of an enrollment increase. Conversely, if training capacity and reimbursement were to keep pace, this mechanism might not operate. The paradox of expansion is a hypothesis about incentive structure, and whether its premises hold must be confirmed with future data.

**Table 2.** Illustrative values used in the expected-value thought experiment.

| Parameter | Definition | Baseline | After expansion | Direction |
| --- | --- | --- | --- | --- |
| P_success | Probability of completing a practice career without a suit-triggering event | 0.90 | 0.85 | Falls if training quality declines |
| R_net | Lifetime net return (normalized to 1 at baseline) | 1.00 | 0.75 | Diluted under a fixed budget |
| P_suit | Expected litigation burden over a practice lifetime | 0.10 | 0.16 | Rises with medico-legal risk |
| C_risk | Cost of a realized suit or loss of license | 5.0 | 5.0 | Determined by law, not by supply |

$E(V) = (P_{\text{success}} \times R_{\text{net}}) - (P_{\text{suit}} \times C_{\text{risk}})$ . Baseline +0.40; after expansion with litigation risk held constant +0.14; after expansion with litigation risk raised -0.16. All values are assumptions adopted for discussion. They are not estimated from observed data and do not predict actual decision-making. They are presented to show which term is moved by policy, and no empirical meaning should be attached to the numbers themselves.

**Figure 2.**
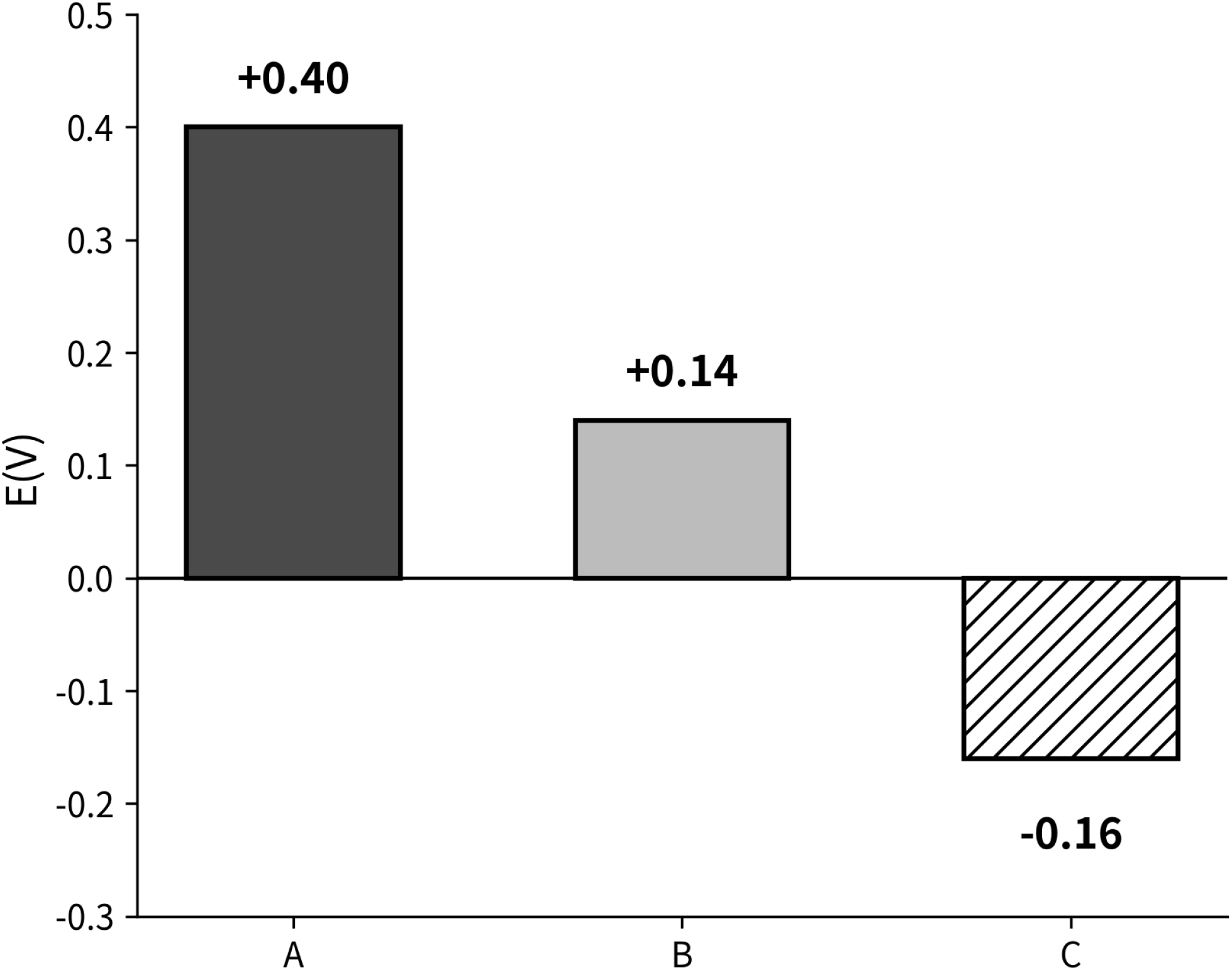
Three illustrative scenarios from the expected-value thought experiment, using the values in Table 2. (A) Baseline. (B) Post-expansion scenario with litigation exposure held constant, in which E(V) declines but does not change sign. (C) Post-expansion scenario with litigation exposure assumed to rise, in which E(V) becomes negative. The sign reversal depends on the assumed link between expansion and medico-legal risk, not on income dilution alone. All values are assumptions adopted for discussion and are not empirical estimates.

As more specialists leave their trained field, the share of already invested training resources that is not realized as essential-care capacity grows with them. Converting that magnitude into a monetary figure, however, would require fixing the social cost invested in training one specialist, the duration of specialty attrition, and the value generated by practice outside the specialty, none of which is supported by published data. This article therefore presents no aggregate estimate. What should be emphasized is that this loss arises among those who have already completed training rather than among those not yet recruited, and that in this sense the target of quota adjustment and the point at which the loss occurs are misaligned.

Emergency medicine is not included in the specialty mismatch data, but it is a case in which the same mechanism can be examined. One study searched criminal court decisions related to emergency care from 2012 to 2021 and identified 22 cases in which physicians were brought to criminal trial on charges of professional negligence resulting in death or injury in the course of emergency care [10]. What is notable here is not the outcome of the final judgment but the number of times the process itself occurred. All 22 cases went through investigation and trial, and in the 12 cases that ended in acquittal the interruption of work and the legal costs incurred along the way arose all the same. This is why P_suit was defined earlier not as the probability of losing an individual case but as the expected burden of litigation over a practice lifetime. Over the same period, declining resident applications to this field and cases of emergency departments unable to accept patients have also been discussed. These data, however, analyze court decisions and do not establish temporal ordering or causation with respect to specialty choice, and emergency care is a domain in which staffing, transfer systems, and bed operations act together. The point here is only that avoidance is observed alongside a large cost term, which is consistent with the expected-value perspective; it is not offered as evidence for it.

Because this argument is conceptual, it is right to state what would count against it. On the expected-value view, if reimbursement and legal risk in an essential field were improved, participation should be expected to recover even without an increase in graduates. Fields of comparable acuity but differing legal exposure should differ in retention. And an enrollment increase implemented while the incentive structure is left unchanged should raise the total number of physicians without raising the workforce in essential fields. If, conversely, essential-field participation rose in proportion to graduate numbers regardless of the return and risk in each field, or if participation did not change after reimbursement was improved and legal exposure reduced, this view would be difficult to sustain. The claim is testable in principle, and the patterns observed so far—the gradient across specialties, and the persistence of vacancy while the physician workforce grew—are not inconsistent with it. This is not evidence confirming the validity of the view, however, but an indication that it is a hypothesis worth examining with appropriate data.

## Conclusion

If participation in essential specialties is more sensitive to the incentives each field offers than to physician supply, the implication runs opposite to the intuition behind expansion. If admissions alone are increased while the expected value of essential practice remains low, it is difficult to expect that the added workforce will necessarily flow into those fields, and if new graduates continue to route around high-risk fields their number may not increase [11]. Conversely, raising the return to essential work and lowering its medico-legal cost so as to restore expected value is what creates the conditions for specialists to re-enter those fields, and in principle this could occur without relying on an enrollment increase. The view that quota adjustment alone can resolve avoidance of essential specialties may not sufficiently account for this layer of incentives. This judgment, however, is derived from the perspective adopted in this article, and its validity should be examined with future data.

This article does not compare specific policy alternatives or estimate their outcomes, and should not be read as doing so. What it proposes is a re-examination of the reasoning that underpins current policy. If the essential-care crisis is viewed not as a shortage of people to train but as a shortage of reasons to stay in those fields, then quota adjustment alone may not readily increase the number of physicians in the specialties that are actually lacking. The conclusion of this article is that a discussion addressing the incentive structure, including reimbursement and medico-legal risk, is needed.

## Data Availability

All data produced in the present work are contained in the manuscript.

## Data availability

Not applicable. All data referred to in this article are from published sources cited in the References.

